# Wildfire Smoke Exposure is Associated with Decreased Sperm Concentration and Total Motile Sperm Count

**DOI:** 10.1101/2025.03.09.25323436

**Authors:** Lillian X. Lindell, Sarah K. Holt, Erin Petersen, Navya Gunaje, Arash Amighi, Amanda Haack, Anthony Bui, Ryan Nasseri, Theodore Crisostomo-Wynne, Catherine J. Karr, Charles H. Muller, Thomas J. Walsh, Tristan M. Nicholson

## Abstract

**Objective:** As wildfires become more prevalent, their impact on fertility warrants evaluation. We aimed to examine the impact of smoke exposure on semen analysis parameters of intrauterine insemination patients in the greater Seattle, WA area. We hypothesized that wildfire smoke exposure was associated with a decline in total motile sperm count.

**Design:** Retrospective cohort study

**Subjects:** Patients undergoing fertility treatments at the University of Washington in 2018-2022

**Exposure:** Subjects were exposed to seasonal wildfire events in the fall of 2018, 2020, and 2022. Pre-exposure semen was a diagnostic fresh sample prior to each respective wildfire event while post-exposure semen was taken at time of intrauterine insemination (IUI) during the wildfire smoke exposure windows. All subjects acted as their own controls in a paired pre-post analysis.

**Main Outcome Measures:** Primary outcome measure was total motile sperm count; secondary outcomes measures are semen volume, sperm concentration, total sperm count, total progressively motile sperm count, percent motile sperm, percent progressively motile sperm.

**Results:** We identified 84 subjects who underwent IUI across the 2018 (n = 27), 2020 (n = 30), and 2022 (n = 27) wildfire smoke events. Median time between initial semen analysis and semen analysis for IUI was 4 months. We observed a decline in sperm concentration, total sperm count, total motile sperm count, and total progressively motile sperm count. We also observed an increase in percent progressively motile sperm. These trends did not differ across event year.

**Conclusions:** Our results are consistent with a prior small study demonstrating that wildfire smoke exposure is associated with declines in sperm quality. These findings highlight the need for further research on the effects of wildfire smoke exposure on human sperm and fertility treatments, especially as smoke exposures are expected to increase with climate change.

## Introduction

Wildfires in the last decade have been increasingly devastating, burning millions of acres and resulting in high intensity of hazardous smoke exposure for days to weeks on end.^1^ The western United States is disproportionately affected by wildfire smoke, with states like California and Oregon experiencing seasonal exacerbations regularly.^1,2^ The Puget Sound region provides an excellent setting for the study of wildfire smoke exposure due to its close proximity to seasonal wildfire activity, and allows researchers to examine the effects of short-term air quality disruptions within a context of otherwise clean air.^3,4^ As wildfire events become more commonplace due to climate change,^5^ it is imperative to understand the effects of their resultant smoke exposure on human health. The impact of wildfire smoke on men’s reproductive health and semen parameters are not well-described and warrant further study, especially as infertility increases in prevalence. This study leverages our institution’s location in the Puget Sound region where wildfire smoke events create distinct pre- and post-exposure periods, allowing for a natural experiment to assess how sudden, temporary surges in wildfire smoke affect semen parameters.

The reproductive health impacts due to air pollution from various sources including fossil fuels, agriculture, and industrial activities are well-characterized. Air pollution of this kind is generally composed of coarse particulate matter (PM) with diameters larger than 2.5 microns (µm),^6^ and is linked to reduced sperm concentration, motility, and morphology.^7,8^ Air pollution has also been linked to declines in conception and adverse pregnancy outcomes, with exposure associated with declines in sperm quality, ovarian dysfunction, and inferior fertility treatments results.^7^

In contrast, PM_2.5_, which are 2.5 µm in diameter or less, is a major component of wildfire smoke.^6^ The smallest particles, ultrafine particulates, defined as compounds less than 0.1 µm in diameter, have been found to make up 30 to 90 percent of wildfire smoke.^9^ Due to the small size of these particulates, they easily diffuse through buildings and affect both indoor and outdoor air quality, making wildfire smoke exposure particularly difficult to avoid.^10^ Therefore, lengthier periods of wildfire smoke exposure can lead to high, sustained levels of fine particulate PM_2.5_ levels both outdoors and indoors.^11,12^

Exposure to PM_2.5_ has been linked to respiratory problems, heart attack, stroke, lung cancer, and cognitive impairment.^13^ However, the impact of PM_2.5_ and wildfire smoke exposure on human male fertility has not been well-characterized in the scientific literature. In mouse models, there is evidence that exposure to wildfire smoke is associated with decreased spermatogenesis and increased sperm DNA methylation, which is a marker for overall male reproductive health.^14,15^ One small cohort study in humans observed that the resultant smoke exposure from the September 2020 wildfire event was associated with decreases in total motile sperm count and sperm concentration in men undergoing intrauterine insemination (IUI) in Portland, OR.^16^ In another retrospective analysis, exposure to high concentrations of PM_2.5_ was associated with an increased odds of azoospermia and lower total progressive motile sperm count.^17^

As wildfire activity increases, it is critical to understand how exposure affects human sperm quality. One in six couples in the world face infertility, with male-factor contributing to approximately 50% of these cases.^18^ Fertility treatments such as IUI and other assisted reproductive technologies (ART) are responsible for a growing share of live births in the United States.^19^ The effect of environmental exposures on sperm viability and reproductive outcomes could have significant impacts on patient counseling and timing of intervention.

We hypothesized that acute hazardous levels of smoke exposure would be associated with a decline in total motile sperm (TMC) in our patient population. Our study seeks to contribute to a growing body of scientific literature on the health impacts of wildfire smoke exposure. Our location in the Puget Sound region offers a unique opportunity to study this phenomenon through a natural experiment. Understanding how wildfire smoke exposure affects semen parameters would impact how patients are counseled prior to fertility interventions, and stimulate future studies on mitigating environmental toxicant exposures in these settings.

## Methods

This retrospective, Institutional Review Board–approved cohort study analyzed diagnostic semen analysis (SA) and SA at time of IUI completed at our institution. We evaluated the last seven years of publicly available air quality data collected by a network of over 60 monitoring stations sponsored by the United States Environmental Protection Agency (EPA), the Washington Department of Ecology, and local Washington clean air agencies.^20^ We defined a smoke event as any period lasting three days above the Washington Air Quality Advisory (WAQA) threshold for fine particulates (PM_2.5_ > 20.4 µg/m³) at the EPA monitoring station closest to our clinic.^21^ We defined a smoke exposure window as a three-month period starting on the first day of the smoke event. We chose to use a three-month window because the process of spermatogenesis takes, on average, 74 days to complete with an additional 14 days of epididymal storage and maturation time prior to emission.^22,23^

Subjects acted as their own controls. We included subjects who had at least one IUI performed during a smoke exposure window, at least one SA performed prior to the first day of the smoke event, and those who lived in the surrounding Puget Sound region during the smoke events. We included individuals’ first IUI that occurred the smoke exposure window, although this was not necessarily their initial IUI. For subjects with multiple SAs before or multiple IUIs during the smoke exposure window, we compared SA temporally closest to the smoke exposure window with the first SA for IUI following the onset of the smoke event. Medical records review was performed to obtain subject age, race, ethnicity, smoking status, marijuana use, infertility history, and illness status at time of IUI. Average amount of smoke exposure and number of smoke days, defined as any day with PM _5_ > 20.4 µg/m³ per the WAQA threshold, were tabulated for all patients at time of SA for IUI based on home zipcodes and the Childs Model (Stanford, CA) of daily smoke PM_2.5_ concentration estimates based on a combination of surface air quality sensor measurements, satellite smoke coverage analysis, and machine learning.^1,2^ Female partner data including fertility history and relevant medical diagnoses were also collected. Our primary outcome was total motile sperm count (TMC), assessed before and after the onset of the smoke exposure. Our secondary outcomes included semen volume, sperm concentration, percent motility, percent progressive motility, total motile sperm, and total progressively motile sperm.

Semen samples for SA were collected and processed following the World Health Organization (WHO) 5^th^ edition criteria for 2018 and 2020 data and 6^th^ edition criteria for 2022 data^24,25^ with the following modifications: Sperm concentration and total sperm count were obtained with use of volumetric measures of semen and hemacytometer counts of sperm in semen diluted with purified water. Sperm motility was evaluated in semen on both cover-slipped wet mounts and Leja Standard Count 20 micron 2 chamber fixed coverslip slides (Leja Products BV, Netherlands). A motility category “Rapid & Linear,” corresponding to the WHO 6th “Rapid Progressive” motility and motility category A of WHO editions prior to the 5th edition was evaluated and recorded for all patients. The diagnostic SA and the SA for IUI were identical, except that morphology and immature germ cell evaluations were only performed for the diagnostic SA. Study data were maintained in a Research Electronic Data Capture (REDCap) database.^26^

Statistical analyses were conducted using R Studio version 4.4.1 (Auckland, NZ) and *RStudio IDE* (Boston, MA). Linear mixed-effects models were employed to evaluate the impact of wildfire smoke exposure on semen parameters including sperm volume, concentration, motility, and count. The number of days between peak wildfire-related PM_2.5_ exposure and semen analysis was also calculated. This variable was used as a covariate in separate time sensitivity analyses to assess whether the timing of IUI relative to the exposure event influenced outcomes. “lme4” package was used to fit models with semen parameters before and after smoke exposure as dependent variables. Models included random intercepts both for patient and wildfire event year to account for temporal variations in the severity of smoke exposure. *p*-Values were determined under the assumption of a two-tailed distribution, with <0.05 considered statistically significant. Results were reported as estimated marginal means.

We identified and included three wildfire events from 2018, 2020, and 2022 as they met our definition of a smoke event as described previously (Figure 1). The 2018 smoke exposure window was from August 14, 2018 to November 14, 2018 (Figure 1A). Peak PM_2.5_ occurred on August 21, 2018 at 102.2 µg/m^3^. The smoke event lasted for 10 days until August 25, 2018, before returning to baseline below 20.4 µg/m^3^ (Figure 1A). The 2020 smoke exposure window was from September 9, 2020 to December 9, 2020 (Figure 1B). Peak PM_2.5_ occurred on September 15, 2020 at 177.7 µg/m^3^. The smoke event lasted for 9 days until September 18, 2020, before returning to baseline (Figure 1B). The 2022 smoke exposure window was from October 7, 2022 to January 7, 2023. Peak PM_2.5_ occurred on October 20, 2022 at 108.0 µg/m^3^. The smoke event lasted for 14 days before returning to baseline on October 21, 2022 (Figure 1C). Notably in 2022, there was a period of unhealthy air from September 10 to September 12, where the average daily PM_2.5_ ranged from 22.2 to 44.5 µg/m^3^. Although this period does not qualify as a smoke event by our definition, the air quality was not in the normal range; therefore, individuals with SAs completed from September 10 to October 7, 2022 were excluded from analysis.

**Figure 1.**
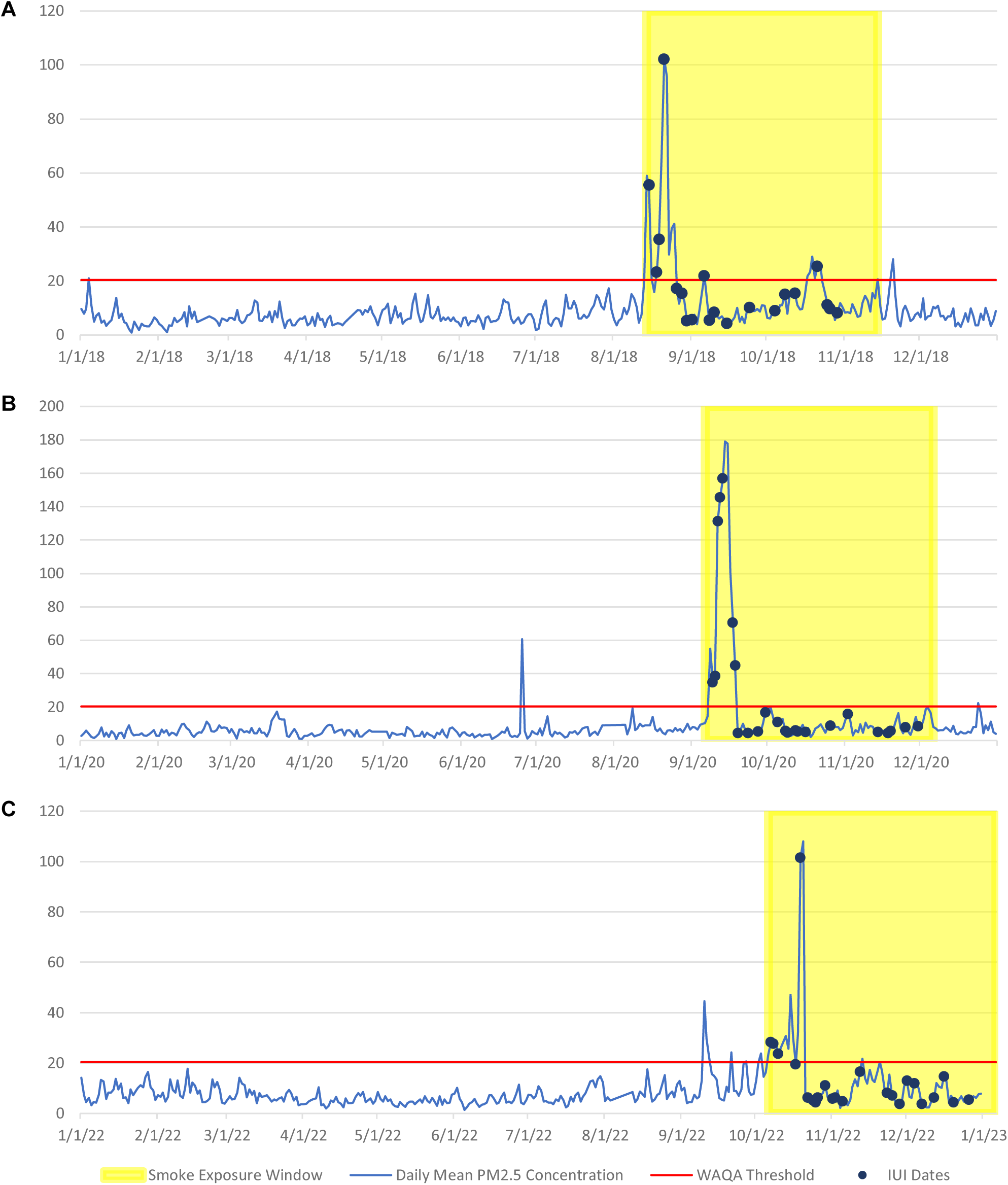
Daily Mean PM_2.5_ in Seattle WA. Daily mean PM2.5 concentration (blue line) as documented by the 10th & Weller Monitoring Station closest to our Men’s Health Lab and Clinic in the years 2018 (**A**), 2020 (**B**), and 2022 (**C**). The red line indicates the Washington Air Quality Advisory (WAQA) threshold for unhealthy air of 20.4 μg/m3, the area highlighted in yellow indicates the three-month wildfire smoke exposure window, and the blue dots represent intrauterine insemination (IUI) dates of included individuals.

## Results

We identified 84 subjects who underwent IUI during the wildfire smoke exposure windows in 2018 (27 subjects), 2020 (30 subjects), and 2022 (27 subjects). Average age at IUI was 37 ± 8 years, with a median time between initial SA and SA for IUI of 4 (3-9) months. 96% of men did not have changes in their fertility history between initial SA and SA for IUI; one individual discontinued a clomid trial, one patient underwent right varicocelectomy, and one patient was recommended over-the-counter CoEnzyme Q10 supplementation, although it is not known if he began taking it in the interim. Median time from day of peak PM_2.5_ to SA for IUI was 17 (1 - 69) days in 2018, 18.5 (1 - 76) days in 2020, and 14.5 (1 - 67) days in 2022. Mean daily PM_2.5_ wildfire smoke exposure for patients in the 90 days preceding IUI was 6.4 µg/m^3^ in 2018, 9.4 µg/m^3^ in 2020, and 6.2 µg/m^3^ in 2022. Mean number of smoke days experienced by patients in the 90 days preceding IUI was 8 ± 2 days in 2018, 9 ± 3 days in 2020, and 7 ± 3 days in 2022.

Most patients were non-Hispanic white individuals with average BMI of 28 ± 5. Average Yost Score of socioeconomic status was 25, with 1 being the highest socioeconomic status and 100 being the lowest.^27^ History taken at time of IUI demonstrated that most men were non-smokers, who had never used marijuana, and had intermittent alcohol intake (Table 1). 69% of men had primary infertility at the time of IUI, and 24% of men were evaluated by our urology practice prior to IUI. The average age of those evaluated by a urologist was 40 ± 13 years-old, with infertility being the most common reason for evaluation (Table 1). The most common infertility-related diagnosis was erectile dysfunction.

**Table 1.** Demographics data of included subjects.

| Parameter | All Cohorts | 2018 | 2020 | 2022 |
| --- | --- | --- | --- | --- |
| Total number of individuals | 84 | 27 | 30 | 27 |
| Average age at IUI (years) | 37 ± 8 | 34 ± 6 | 36 ± 4 | 40 ± 12 |
| Median time between SA and IUI (months) | 4 (3 – 9) | 3 (2.5 – 5) | 4 (2 – 9) | 5 (3 – 10) |
| Average BMI | 28 ± 5 | 27 ± 4 | 29 ± 6 | 27 ± 6 |
| Average Yost Score of socioeconomic status | 25 ± 22 | 21 ± 21 | 20 ± 17 | 31 ± 24 |
| Census Bureau race group |  |  |  |  |
| White | 41 (49%) | 12 (44%) | 17 (57%) | 12 (44%) |
| Black or African American | 6 (7%) | 2 (7%) | 2 (7%) | 2 (7%) |
| Asian | 26 (31%) | 11 (41%) | 5 (17%) | 10 (37%) |
| Native American or Alaskan Native | 1 (1%) | 0 (0%) | 1 (3%) | 0 (0%) |
| Declined to answer or unknown | 10 (12%) | 2 (7%) | 5 (17%) | 3 (11%) |
| Ethnicity |  |  |  |  |
| Not Hispanic, Latino, or Spanish | 71 (85%) | 23 (85%) | 25 (83%) | 23 (85%) |
| Hispanic, Latino, or Spanish | 3 (4%) | 1 (4%) | 0 (0%) | 2 (7%) |
| Declined to answer or unknown | 10 (12%) | 3 (11%) | 5 (17%) | 2 (7%) |
| Smoking History |  |  |  |  |
| Never | 65 (77%) | 21 (8%) | 24 (80%) | 20 (74%) |
| Former | 13 (15%) | 3 (11%) | 4 (13%) | 6 (22%) |
| Current | 4 (5%) | 3 (11%) | 0 (0%) | 1 (4%) |
| Unknown | 2 (2%) | 0 (0%) | 2 (7%) | 0 (0%) |
| Marijuana History |  |  |  |  |
| Never | 50 (60%) | 20 (74%) | 15 (50%) | 15 (56%) |
| Former | 14 (17%) | 4 (15%) | 5 (17%) | 5 (19%) |
| Current | 17 (20%) | 3 (11%) | 7 (23%) | 7 (26%) |
| Unknown | 3 (4%) | 0 (0%) | 3 (10%) | 0 (0%) |
| Alcohol History |  |  |  |  |
| None | 20 (24%) | 10 (37%) | 4 (13%) | 6 (22%) |
| Social (intermittent) | 50 (60%) | 11 (41%) | 20 (67%) | 19 (70%) |
| Regular (daily) | 3 (4%) | 2 (7%) | 1 (3%) | 0 (0%) |
| Unknown | 11 (13%) | 4 (15%) | 5 (17%) | 2 (2%) |
| Male Infertility Type |  |  |  |  |
| Primary Infertility | 58 (69%) | 19 (70%) | 23 (77%) | 16 (60%) |
| Secondary Infertility | 26 (31%) | 8 (30%) | 7 (23%) | 11 (40%) |
| Infertility-Related Diagnoses |  |  |  |  |
| Solitary testicle | 1 (1%) | 1 (4%) | 0 (0%) | 0 (0%) |
| Erectile dysfunction | 5 (6%) | 1 (4%) | 1 (3%) | 3 (11%) |
| HIV | 1 (1%) | 0 (0%) | 0 (0%) | 1 (4%) |
| Pituitary micro/macroadenoma | 1 (1%) | 1 (4%) | 0 (0%) | 0 (0%) |
| Prior testosterone use | 1 (1%) | 0 (0%) | 0 (0%) | 1 (4%) |
| Radiation exposure | 2 (2%) | 1 (4%) | 1 (3%) | 0 (0%) |
| Elevated scrotal temperatures (regular hot tub use) | 7 (8%) | 2 (8%) | 4 (13%) | 1 (4%) |
| Pesticides or other organic compounds | 2 (2%) | 0 (0%) | 0 (0%) | 2 (8%) |
| Febrile Illness within prior 3 months of IUI |  |  |  |  |
| Yes | 3 (4%) | 1 (4%) | 1 (3%) | 1 (4%) |
| No | 77 (92%) | 24 (89%) | 29 (97%) | 24 (89%) |
| Unknown | 4 (5%) | 2 (7%) | 0 (0%) | 2 (7%) |
| Number of men evaluated by urology prior to IUI | 24 (29%) | 7 (26%) | 6 (20%) | 11 (41%) |
| Average age of men evaluated by urology (years) | 40 ± 13 | 34 ± 3 | 39 ± 4 | 45 ± 18 |
| Reasons for initial urologic evaluation (multiple selection) |  |  |  |  |
| Infertility evaluation | 20 (83%) | 6 (86%) | 6 (100%) | 8 (73%) |
| Fertility preservation | 1 (4%) | 0 (0%) | 0 (0%) | 1 (9%) |
| History of vasectomy, desire for restoration of fertility | 1 (4%) | 0 (0%) | 0 (0%) | 1 (9%) |
| Varicocele | 1 (4%) | 1 (14%) | 0 (0%) | 0 (0%) |
| Testicular or pelvic pain | 1 (4%) | 1 (14%) | 0 (0%) | 0 (0%) |
| Low testosterone | 1 (4%) | 0 (0%) | 0 (0%) | 1 (9%) |
| Erectile dysfunction | 1 (4%) | 0 (0%) | 1 (17%) | 0 (0%) |
| Ejaculatory Dysfunction | 1 (4%) | 0 (0%) | 1 (17%) | 0 (0%) |

Smoke exposure was estimated to correlate with a decline of 133 million total sperm (*p* = 2.3 x 10^-5^) and 71 million total motile sperm (*p* = 2.4 x 10^-5^, Table 2). Semen volume also declined by an estimated 0.6mL (*p* = 1.6 x 10^-6^) and sperm concentration by 22 million/mL (*p* = 2.2 x 10^-3^, Table 2). In our linear mixed-effect models, event year and time from peak PM_2.5_ exposure as covariates did not have statistically significant impacts on semen parameters. All *p*-values for event year effect and time from peak exposure were not statistically significant.

**Table 2.** Estimated effect of wildfire smoke on median semen parameters of 84 subjects before and after exposure.

| Semen Parameters | Before Exposure | After Exposure | Estimated Effect* | P value** |
| --- | --- | --- | --- | --- |
| Semen volume (mL) | 3.5 (2.3 – 4.3) | 3.1 (2.3 – 3.7) | -0.6 | **1.6 x 10<sup>-6</sup>** |
| Sperm concentration (mil/mL) | 76 (49 – 127) | 74 (33 – 102) | -22 | **2.2 x 10<sup>-3</sup>** |
| Percent motile (%) | 54 (47 – 60) | 53 (44 – 62) | -0.4 | 0.800 |
| Percent progressively motile (%) | 39 (32 – 66) | 46 (38 – 56) | 7 | **4.4 x 10<sup>-5</sup>** |
| Percent rapid & linear motile (%) | 13.5 (10 – 18) | 12.5 (5.5 – 18) | -2 | 0.055 |
| Total sperm count (x10<sup>6</sup>) | 286 (177 – 419) | 184 (98 – 332) | -133 | **2.3 x 10<sup>-5</sup>** |
| Total motile sperm (x10<sup>6</sup>) | 157 (80 – 250) | 90 (35 – 188) | -71 | **2.4 x 10<sup>-5</sup>** |
| Total progressively motile sperm (x10<sup>6</sup>) | 106 (46 – 175) | 77 (38 – 158) | -37 | **4.9 x 10<sup>-3</sup>** |
| Total rapid & linear motile sperm (x10<sup>6</sup>) | 34 (15 – 79) | 20 (7 – 60) | -22 | **1.2 x 10<sup>-4</sup>** |

\*Linear mixed model estimate of the effect of wildfire smoke exposure when controlling for event year
\*\*Linear mixed model evaluation with $p < 0.05$ considered statistically significant, event year nor time from peak wildfire exposure did not have statistically significant impact for any independent variables so p-values are not reported

Partner data and observed reproductive outcomes were also summarized (Table 3). Of the 84 female partners, average age was 34 ± 5 years at time of IUI. The majority of these women had never been pregnant. All partners were evaluated by reproductive endocrinologists, and no female factor infertility was identified in 36% of cases following workup. Two semen samples for IUI were ultimately not inseminated due to low sperm count, and one sample was taken to an outside institution for insemination. Of the remaining 81 IUIs, 9 (11%) resulted in pregnancy and 7 (9%) resulted in live birth with average gestational age of 39 ± 3 weeks at birth (Table 3).

**Table 3.** Partner data and reproductive outcomes.

| Parameter | All Cohorts |
| --- | --- |
| Average female partner age at IUI (years) | 34 ± 5 |
| Median number of pregnancies prior |  |
| Gravida | 0 (0 – 1) |
| Para | 0 (0 – 0) |
| Abortus | 0 (0 – 1) |
| Partner female factory infertility history |  |
| No female factor noted after workup | 30 (36%) |
| Tubal factors | 5 (6%) |
| Uterine factors | 10 (12%) |
| Cervical factors | 1 (1%) |
| Diminished Ovarian Reserve | 12 (14%) |
| Polycystic Ovarian Syndrome | 20 (24%) |
| Endometriosis | 4 (5%) |
| Other | 17 (20%) |
| Number of non-inseminated samples due to low count | 2 |
| Number of pregnancies achieved from IUI | 9 (11%) |
| <35 years-old at conception | 5 (56%) |
| 35-40 years-old at conception | 4 (44%) |
| >40 years-old at conception | 0 (0%) |
| Number of live births | 7 (9%) |
| <35 years-old at birth | 3 (43%) |
| 35-40 years-old at birth | 4 (57%) |
| >40 years-old at birth | 0 (0%) |
| Average gestational age at birth (weeks) | 39 ± 3 |

## Discussion

The increasing frequency and intensity of wildfires highlight the need to understand how poor air quality and fine particulate pollution affect fertility. In our study, patients undergoing IUI treatment had significant declines in numerous semen parameters after wildfire smoke exposure. We found that wildfire smoke exposure was associated with declines in semen volume, sperm concentration, total sperm count, total motile sperm count, total progressively motile sperm count, and total rapid and linear motile sperm count (Table 2).

There was also a statistically significant increase in percent progressively motile sperm post-exposure. Our findings are similar to those in a smaller cohort study examining changes in semen parameters after the 2020 wildfire event,^16^ and corroborate a larger analysis that associates high PM_2.5_ levels with increased odds of lower total progressively motile sperm count.^17^ Together, these findings support our hypothesis that wildfire smoke exposure is associated with declines in sperm parameters.

Spermatogenesis occurs in cyclical waves over an average of 74 days.^22^ Additionally, fully-formed sperm undergo maturation and storage in the epididymis for approximately two weeks prior to ejaculation.^23^ It is unclear at which points during this process spermatozoa and sperm are vulnerable to environmental toxicant exposures such as wildfire smoke. Although individuals in our cohort were completing their SAs for IUI throughout the smoke exposure period, the median absolute time from exposure to SA each year was 14-19 days. One plausible pathway for this rapid impact of wildfire smoke exposure on semen parameters is through direct toxicant effect on mature sperm during epididymal storage or transit through the vas deferens, as sperm are particularly susceptible to noxious insults due to the lack of DNA repair mechanisms in the epididymis.^28^ Oxidative stress induced by inhaled pollutants have been shown to damage sperm membranes and DNA rapidly, even in the absence of effects on spermatogenesis.^29^ Additionally, toxicants may disrupt epididymal function, which may impair sperm maturation and may lead to changes in semen quality within a few weeks.^30^ It is also possible that toxicity during a specific stage of spermatogenesis may underlie our findings.

However, addressing this latter hypothesis will likely require large-scale epidemiological studies with greater sample sizes and temporal resolution, or targeted investigations using animal models.

We also observed a statistically significant rise in the percentage of progressively motile sperm after wildfire exposure (Table 2). One possible theory for this observation is that abnormally motile sperm are more vulnerable to the oxidative stress imparted by inhaled fine particulates. Sperm progressive motility has been found to correlate positively with DNA integrity, whereas higher levels of DNA fragmentation are found in poorly motile sperm.^31^

Sperm with higher levels of baseline DNA fragmentation, as evidenced by poor motility, may be more susceptible to culling during interaction with acute stressors like fine particulate wildfire smoke. These findings highlight the need for additional research on the effects of wildfire smoke exposure on vulnerable populations such as men with elevated levels of sperm DNA fragmentation.

In 2020, the maximum fine particulate PM_2.5_ level in the Seattle area during a regional wildfire event was of 177.7 µg/m^3^, which was less than half of the maximum fine particulate level in the Portland, OR region of PM_2.5_ 388 µg/m^3^.^16^ Importantly, we observed the same magnitude of impairment of semen parameters with a lower PM_2.5_ level when compared to the Portland, OR study that same year. When comparing between events in our Seattle area study, we note that the maximum PM_2.5_ level in 2018 was slightly over half of the PM_2.5_ level in 2020 (102.2 vs 177.7 µg/m^3^, respectively); however, event year, and by association severity of wildfire smoke, did not seem to correlate with worse declines in semen parameters. Whether these findings suggest fine particulate exposure has a threshold effect on changes in semen parameters warrants further research.

While this study was not specifically designed to evaluate the impact of wildfire smoke exposure on reproductive outcomes, our observational partner data highlight areas that merit further investigation. In our cohort of women, whose average age was 34 years at time of IUI, pregnancy rate was 11% and live birth rate was 9%, which are similar to the 12-20% pregnancy rate and 9-15% live birth rate per cycle for women in their mid-thirties reported in the literature.^32–34^ Data collected at our institution from 1995-2017 demonstrate a pregnancy rate of 16% and live birth rate of 6% after IUI.^35^ This underscores the need for future research, as the observed impact on IUI outcomes has important implications for patient counseling and ART procedural timing.

Our study had several strengths. We were able to take advantage of a natural experiment in a region with good air quality outside of wildfire smoke season where subjects could act as their own controls. The data analyzed only included couples undergoing their first IUI during a smoke window, and all our analyses were performed at the same center and by the same staff. We also collected detailed clinical information about our subjects taken at time of IUI, and all collections were done in person. Although we could not measure total time spent outdoors versus indoors for subjects, prolonged unhealthy air quality outdoors is associated with parallel declines in indoor air quality^10,11^ and all individuals in this study traveled to our clinic for semen analysis and IUI during periods of wildfire smoke, inevitably resulting in some level of exposure to poor outdoor air.

Limitations of our study include small sample size and unmeasured variables that may mitigate or exacerbate the health effects of wildfire smoke exposure. These include factors that influence individual exposure such as the estimated duration of outdoor smoke exposure, wearing of high-quality masks, use of indoor air cleaners, occupation, change in behaviors due to wildfire smoke exposure, and exercise habits. Additionally, the subjects in our study live in relatively high socioeconomic regions, which is not representative of the general population. It is also possible that baseline declines in semen analysis parameters occur around the time of IUI due to the inherent stress of the procedure.

Another potential unmeasured influence on sperm quality is the COVID-19 pandemic for 2020 and 2022 cohorts, as COVID-19 infection has been shown to have transient impact on sperm quality.^36,37^ Even mild or asymptomatic COVID infection has been found to decrease sperm concentration, total sperm count, motility, and morphology.^38^ Our intake forms at time of IUI asked patients about recent illness including fever and cough, to which the vast majority of subjects answered “no” (Table 2). However, mandatory COVID testing prior to both diagnostic SA and IUI appointments was not conducted. It is therefore unknown if asymptomatic COVID infection before or after smoke exposure could have had an impact on semen parameter results. However, the likelihood this impacted our results is low, as the semen parameters were not statistically different in the 2018 wildfire event year. Future prospective studies where data are collected on subject behavior and symptoms are required to address these concerns.

Most of the smoke days in the Puget Sound region are also the result of remote wildfires that began in northeast Washington and Oregon. It is well-documented that the chemical composition of wildfire smoke changes over time and space as it travels from its location of origin.^39^ Oxidation of organic compounds and the formation of secondary pollutants such as ozone and peroxyacetyl nitrates may have a distinct effect on semen parameters, which warrant further investigation.

Lastly, the timeline for exposure was not studied and is a critical area for further research. We used a three-month smoke exposure window as a biologically plausible timeframe; however, we do not know if there are more time-resolved sensitive windows of sperm vulnerability from acute smoke exposure, if the effects of exposure could persist beyond three months, and if and when semen parameters eventually recover. Interestingly, when we accounted for time from peak wildfire smoke PM_2.5_ exposure to SA for IUI, we did not see statistically significant effects. However, previous studies have shown that the timing of exposure may influence certain semen parameters. Ramsay et al’s analysis of the Subfertility, Health and Assisted Reproduction (SHARE) data demonstrates that progressively motile sperm count appears most affected by exposure 60-74 days prior to SA, whereas azoospermia was more likely with exposure 30-59 days prior to SA.^17^ These are compelling evidence that wildfire smoke has acute deleterious effects during the process of spermatogenesis, storage, and transport of sperm, and perhaps suggest that cumulative exposure may play a more critical role than the timing relative to peak exposure. Ultimately, the window of exposure relevant for human sperm and fertility is not fully understood and requires further study.

## Conclusions

In this study, we examined the impact of wildfire smoke exposure on male fertility, finding significant declines in several semen parameters, including sperm concentration, total motile sperm count, progressively motile sperm count, and rapid and linear motile sperm count after exposure to fine particulate pollution. These results emphasize the need for further research on the short- and long-term effects of wildfire smoke exposure on reproductive health, especially as wildfire smoke events become more frequent with climate change.

## Data Availability

All data produced in the present study are available upon reasonable request to the authors

