## Supplemental Figure 1 for "Wildfire Smoke Exposure is Associated with Decreased Sperm Concentration and Total Motile Sperm Count"

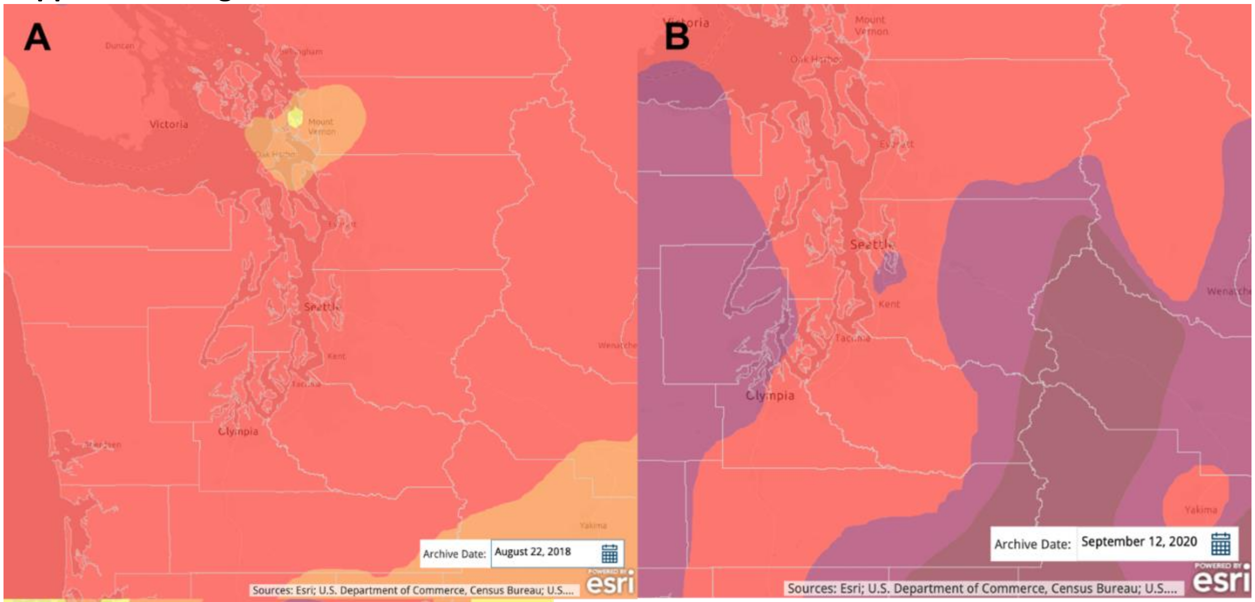


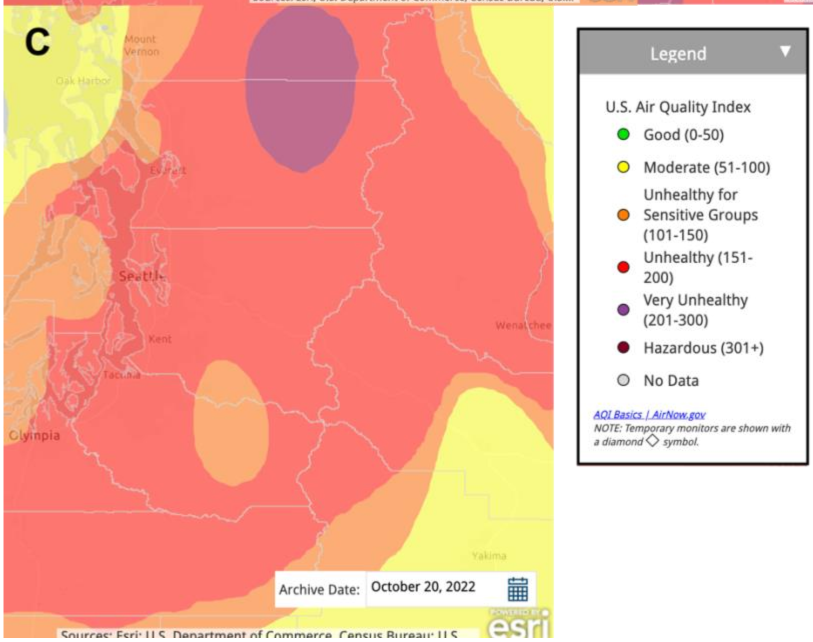


Maps of $PM_{2.5}$ levels on worst day of air during the (**A**) Aug 2018, (**B**) Sept 2020, and (**C**) Oct 2022 smoke events as visualized using ArcGIS Online (Esri, 2025).[40]
Color-coded legend of $PM_{2.5}$ overlay taken from ArcGIS (Esri, 2025).
